# Accelerated Partner Therapy contact tracing intervention for people with chlamydia: the LUSTRUM process evaluation using programme theory

**DOI:** 10.1101/2021.08.07.21261736

**Authors:** Paul Flowers, Fiona Mapp, Jean McQueen, Rak Nandwani, The LUSTRUM programme, Claudia Estcourt

**Affiliations:** School of Psychological Sciences & Health, University of Strathclyde, Glasgow, UK; Institute for Global Health, University College London, London, UK; School of Health and Life Sciences, Glasgow Caledonian University, Glasgow, UK; NHS Greater Glasgow and Clyde, Glasgow, UK

## Abstract

**Background:** Using programme theory we report a process evaluation of Accelerated Partner Therapy (APT) - a novel contact tracing (partner notification) intervention for people with chlamydia as part of the LUSTRUM trial.

**Methods:** Following the specification and visualisation of initial programme theory, questions of context dependency, fidelity, and functioning of putative intervention mechanisms were addressed using deductive thematic analysis of qualitative data collected through focus groups and individual interviews with purposively sampled health care professionals (n=34 from ten sites), index patients (n=15), and sex partners who received APT (n=17). Analyses were independent of trial results.

**Results:** APT was anticipated to change key interactions and sexual health service organisation to accommodate safe and optimal remote care. APT training and resources transformed key interactions as anticipated. Overall intervention fidelity was good and APT was well-liked by those who delivered and received it. Putative intervention mechanisms worked mostly as expected although those concerned with local implementation sometimes worked counter to expectations. APT and its trial struggled to be implemented at scale across all sites. Considerable pressures drove services to constantly adapt to achieve efficiencies. APT was perceived as time consuming without visible impact on perceived patient numbers in clinic curtailing positive feedback loops driving normalisation.

**Discussion:** Using programme theory we show an evidence-based, theoretically informed, overview of how APT worked dynamically within the context of the trial and within UK sexual health services. We find a mixed picture of a well-liked, intuitive, coherent intervention struggling to gain purchase within an already pressured service.

**Trial registration:** <u>ISRCTN15996256</u>

**Study protocol:** <u>doi.org/10.1136/bmjopen-2019-034806</u>

**Ethical approval:** This study received ethical approval from London—Chelsea Research Ethics Committee (18/LO/0773). Findings will be published with open access licences.

**Funding:** This work presents independent research funded by the National Institute for Health Research (NIHR) under its Programme Grants for Applied Research Programme (reference number RP-PG-0614-20009).

## INTRODUCTION

Here we report a process evaluation of a contact tracing intervention within sexual health (i.e. Accelerated Partner Therapy (APT)). Within sexual health contact tracing is typically referred to as partner notification (PN). The LUSTRUM programme (Limiting Undetected Sexually Transmitted infections to RedUce Morbidity, (Estcourt *et al*., 2020) builds on previous work demonstrating APT to be feasible and acceptable (PN) and able to treat higher proportions of sex partners of people initially diagnosed with chlamydia (i.e index patients) than usual care (Estcourt *et al*., 2012, 2015). APT ‘fast-tracks’ testing and treatment for the sexual partners of an index patient (i.e. the person initially diagnosed with the sexually transmitted infection (STI)) by offering a timely telephone consultation with a trained healthcare professional (HCP) who then, if it is safe to do so, arranges rapid delivery of an STI self-sampling and treatment pack to the sex partner via the index patient or by post to be used at home. The LUSTRUM programme included a cluster randomised control trial where each of the 14 sites offered APT vs routine care, or routine care followed by APT.

These interactions are intended to *accelerate* treatment of chlamydia and reduce the likelihood of the STI being passed on; either back and forth between the index patient and their sex partners, or indeed onwards to other people. Within the LUSTRUM programme we optimised these interactions between HCP, index patient and sex partner creating an intervention manual (Pothoulaki *et al*., 2021) and enhanced self-sampling and treatment packs (Flowers *et al*., 2020) that together persuade and enable all concerned to enact APT. Yet these interactional chains are all mediated by the complex context of sexual health service provision. At the time of the LUSTRUM programme such services were not organised to deliver this remote care, or to offer partner-delivered, or postal, treatment. Instead sexual health services are geared towards the inherent uncertainties of infectious disease (i.e. patterns in transmission) and they tend to offer a mixture of drop-in and bookable appointments to accommodate the flow of new patients and the follow up of patients. Within the UK there has been a recent disinvestment within this area of health. In some countries of the UK, sexual health services are no longer commissioned by the National Health Service but instead by local authorities (Robertson *et al*., 2017).

A process evaluation was designed to explore APT within a multi-site cluster randomised controlled trial. We used programme theory as a central device to structure the process evaluation (data collection, analysis and write up). Programme theory is a term used to describe how interventions and their actions can be made explicit and theorised. Interventions have been described as ‘theories incarnate’ (Pawson & Tilley, 1997) given they consolidate and enact prior expertise from various sources including professional intuition and practice, formal theories, and/or common sense. Programme theory is intended to reflect the dynamic flow of an intervention and all its moving parts rather than examine one or two elements of an intervention alone. Its focus on *how* an intervention works rather than *if* an interventions works is intended to provide useful knowledge. Across the literature, key elements of programme theory are variable and there is no single model of what it should consist of but typical content usually describes the context of an intervention and its contextual dependencies (which elements of the context are needed), the specific problem the intervention aims to address, the intervention components (and sometimes their interactions), intervention mechanisms (and sometimes their interactions) and a range of intervention outcomes (e.g., see Baranowski and Stables, 2000; Steckler and Linnan, 2002; Grant and colleagues (2013a); MRC guidance on process evaluation). Programme theory is often written narratively but also visualised within logic models. The latter show an overview of the inherent logic, or causal mechanisms of a given intervention, binding content to outcomes within given contexts.

When used within process evaluation programme theory may be used to compare how an intervention was thought to work *prior* to a trial, compared to how an intervention *actually worked* within a trial. Given the focus of programme theory on the dynamic whole of the intervention rather than any single part of it in isolation, this approach can be particularly useful in considering the transferability and/or sustainability of interventions following some demonstration of efficacy. The approach is less useful for explaining specific trial results where more traditional sequential mixed methods research designs may be more useful (e.g. Mapp et al., 2021a). Here we report our process evaluation of APT conducted before the main trial results were shared with the process evaluation team. We use the structure of our programme theory to consider what we have learned from about APT within the context of the LUSTRUM trial, and to prompt detailed consideration of pertinent issues for the future scale up of APT outwith a trial context, should the trial results show clinical significance.

## RESEARCH QUESTIONS

1. How was APT thought to work as a complex intervention?
2. What did we learn about the context and contextual dependencies?
3. Were the intervention components delivered as intended?
4. Is there evidence to support the causal mechanisms by which the intervention was intended to work?

## METHOD

We used a four-step process to conduct this process evaluation using programme theory

### Step 1: Developing the initial programme theory

Initial programme theory was developed to provide multi-levelled account of *how* we thought APT worked. This was achieved through an iterative combination of documentary analysis, video analysis of APT being delivered, an exploration of the key sequential steps of PN interventions within the published literature (see Mapp et al, 2021b), earlier qualitative work on the psychosocial and health service context of PN (Pothoulaki et al., 2020) and iterative discussions and presentations to the wider trial team. This initial programme theory was visualised in terms of a series of logic models depicting key elements of APT and its contexts and presented and discussed with the whole team until consensus was reached. To theorise *how* we anticipated APT to work at individual and service levels we drew upon a range of approaches including normalisation process theory (Murray et al., 2010), the behaviour change wheel approach (Michie et al., 2011), the theoretical domains framework (Atkins et al., 2017) and concepts from systems science (e.g. Rutter et al., 2017). In this way our initial programme theory provided a hypothetical theorised framework of the way the content affected the outcomes within given contexts.

### Step 2: Collecting data from healthcare professionals, index patients and sex partners involved in APT

We recruited heterogeneous purposive samples of HCPs involved in the trial. Efforts were made to recruit from a range of sites including those that recruited large and small numbers of patients for APT Recruitment sites spanned urban and rural areas in Scotland and England and broadly represented the full range of APT trial sites. A total of 34 healthcare professionals from twelve sites who had been involved in delivering APT as part of the LUSTRUM trial took part in one-off focus groups (HCP FG) (n=6) or one-to-one interviews (1:1) (n=10). Data collection was conducted by PF. We also conducted telephone interviews 15 index patients and 17 sex partners. Data collection was conducted by JMcQ. Semi-structured topic guides were used to collect data that focussed broadly on experiences of APT (see Appendix 1).

### Step 3: Analysing data within the framework of our programme theory

Data were analysed deductively using a relatively structured approach employing a pro-forma to ensure that analysis focussed on the main categories of the programme theory. The analysts were also open to new inductively derived findings being identified within the data however. Analysis of the HCP data was led by PF and analysis of index and sex partner data was led by FM. PF and FM subsequently audited each other’s work and engaged in discussions about the whole analysis before presenting findings to the wider team. These analyses took place before the main trial results were analysed. Analysis did not compare or contrast individual trial sites and the authors were blind to the relative trial activities in each site at the point of data collection and analysis. Equally analysis did not focus on comparing differences between sites that offered APT prior to being control vs being a control and then APT. Instead analysis focussed on building a picture of the general delivery of APT and the trial across time and across all sites. The relative strength of the themes described reflects the frequency and gravity of the data collected both across and within sites. Where relevant we further analysed these deductive thematic analyses with the three complementary lenses used to theorise our initial programme theory; the behaviour change wheel approach (Michie et al., 2011), implementation science (Murray et al., 2010), and complex adaptive systems concepts (e.g. Rutter et al., 2017).

### Step 4: Visualising the revised programme theory through a new logic model

PF colour-coded the findings and the logic model using a Red-Amber-Green traffic light system to provide a high-level overview. These were audited and check by FM before being presented to the wider research team for comment and debate.

This process evaluation study received ethical approval from London—Chelsea Research Ethics Committee (18/LO/0773).

## RESULTS

### Participant characteristics (index patient and sex partners)

We collected data from 15 index patients and 17 sex partners (including six pairs where the sex partner was linked to the index patient) between July and December 2019. All index patients interviewed were female with a mean age of 23 years (age range of 18-32 years). Ten described themselves as White British/White Other ethnicity, two were Black/Black British, two were Asian/Asian British and one index patient was of mixed ethnic background. Index patients reported between one and five sex partners^1^; 13 described themselves as being in a relationship (mostly long-term or dating, one was a short-term casual arrangement). Education levels varied with seven having completed secondary education, four had finished college and four had completed university level education. Two index patient participants were unemployed at the time of interview.

Sex partner participants differed from index patients interviewed as 12 of the 17 were male with a mean age of 25 (age range 18-34 years). Ten reported being in a relationship when interviewed. Three had a secondary education level, seven had finished college and the remaining seven had a university education. One person was still in education and one was unemployed when they were interviewed. We did not collect data on ethnicity for sex partners.

### Research Question 1: How did we anticipate APT working?

A narrative account of this *initial* programme theory is available as a supplementary file (*supplementary file 1*).

Prior to the process data collection and analysis the APT intervention was conceptualised as working simultaneously at different levels. There is the interaction of the key individuals and their *behaviour change* (including health care professionals, index patients and sex partners); the level of sexual health services in developing locally *implementation* plans suited to their resources and local set up; and, at the system level, with APT working simultaneously within a number of complex and adaptive *systems (e*.*g*., *digitization, health care and sexual cultures)*. Thus we anticipated that the behaviour change of key individuals was realised within particular settings, and these in turn were embedded within wider systems.

The programme theory was used to underpin our process evaluation by detailing this multi-levelled complexity and the key processes by which APT was imagined to work. Here within the main body of the paper we illustrate a simplification and visualisation of our initial programme theory by focusing on our final logic model. Figure 1 provides an overview of key aspects of context, the problem APT is attempting to resolve, intervention components, intervention mechanisms and their relationship to the main trial outcomes.

**Figure 1:**
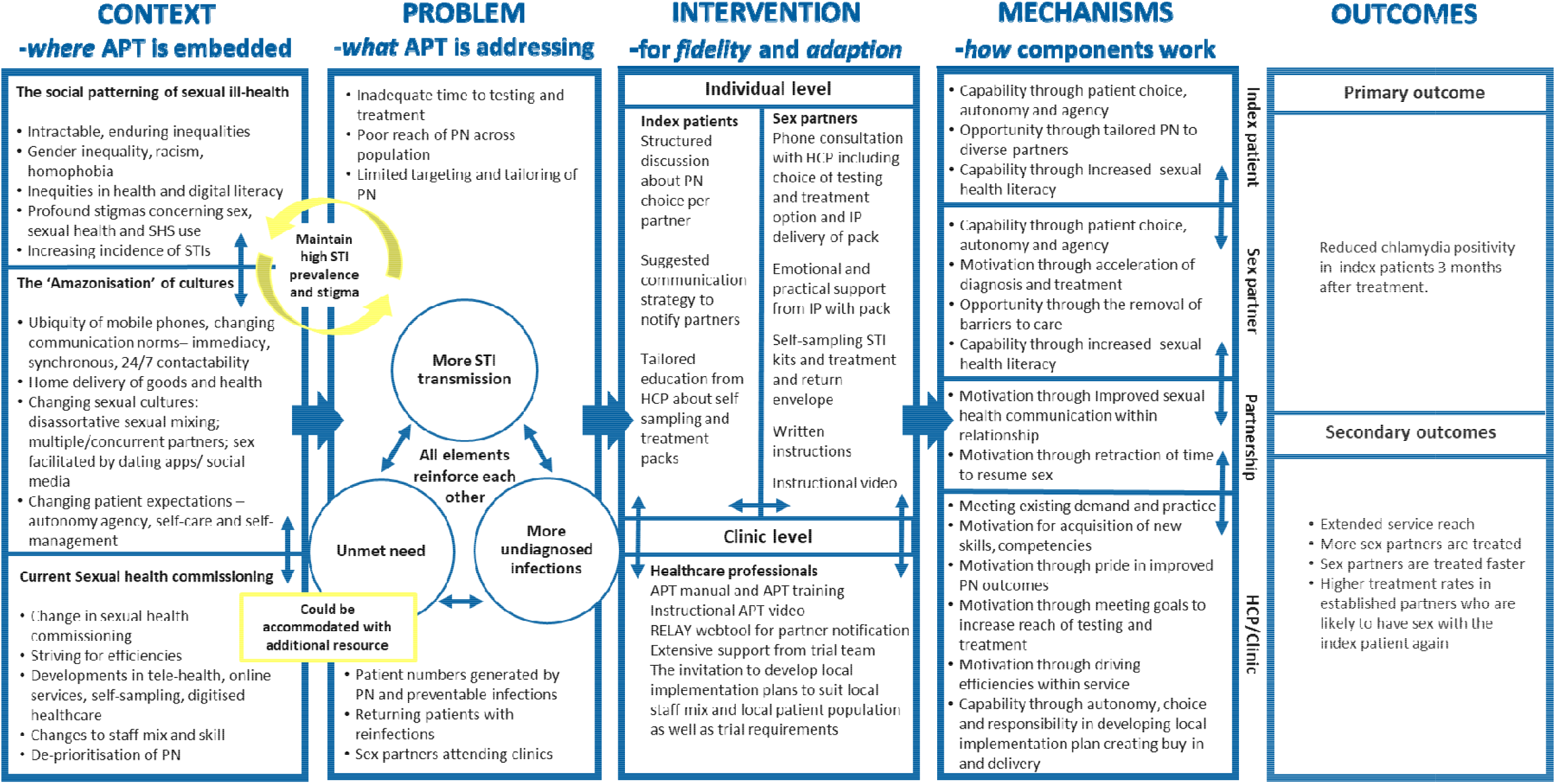
The initial programme theory of Accelerated Partner Therapy.

**Figure 2:**
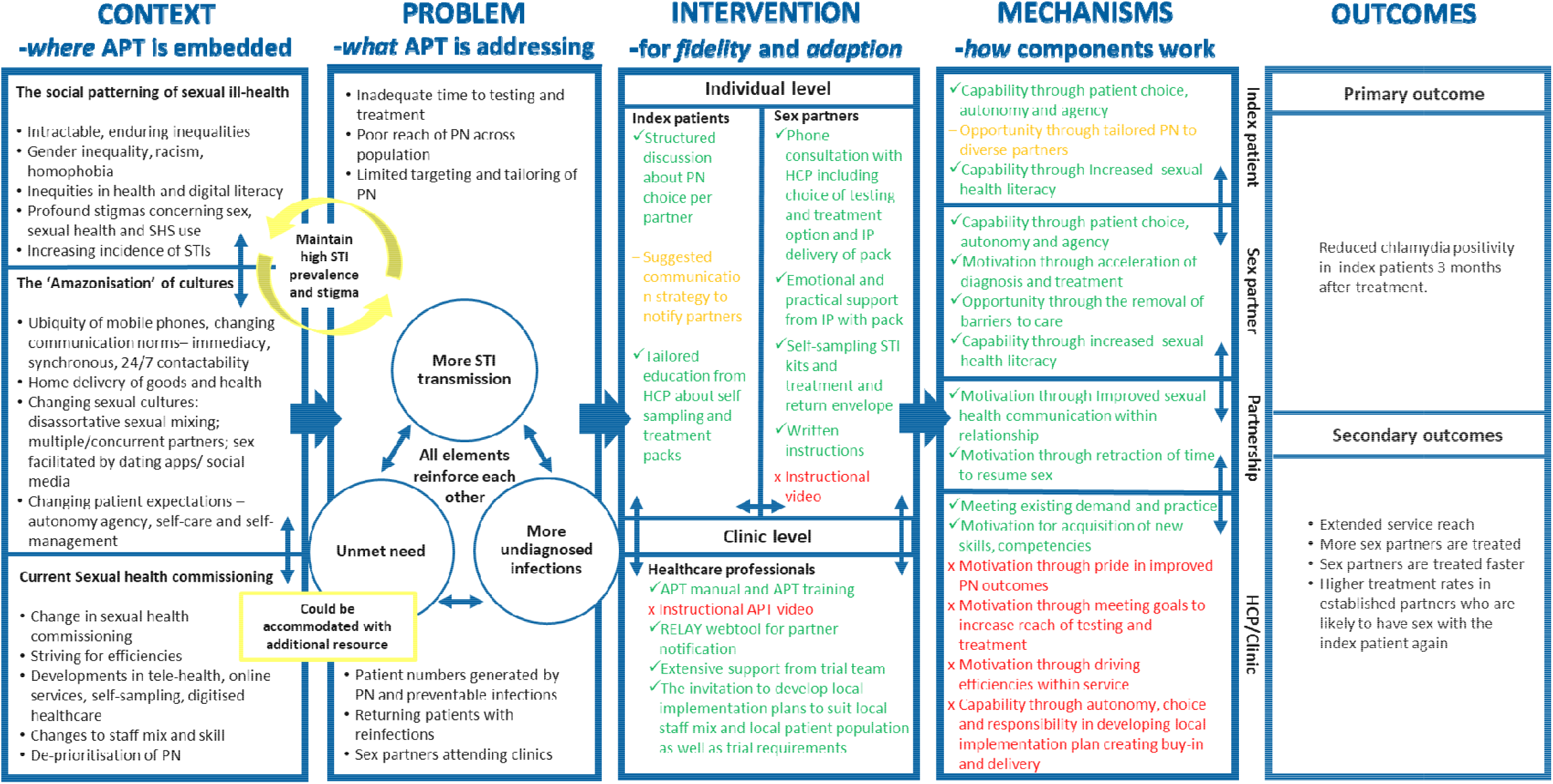
The revised programme theory of Accelerated Partner Therapy.

#### Contexts

It shows a multi-layered understanding of the context highlighting the intersections between different intersecting complex systems, for example, the social patterning of sexual ill health, the ‘amazonisation’ of wider culture associated with digitalisation, and recent historic changes to the structure of sexual health services.

#### The problem APT is attempting to solve

Using concepts from systems science it shows these contextual elements are interlinked and feed into a vicious circle in which reinforcing feedback loops such as increasing numbers of STI infections are associated with decreasing capacity within the system to meet sexual health need which in turn leads to emergent effects such as further onwards transmission of STIs. Although relatively modest in its aspirations, APT attempts to address some aspects of this complexity by rapidly testing and treating people highly likely to have been exposed (i.e., the sex partners of people very recently diagnosed) and thus slowing onwards transmission and disrupting the vicious circle of STI transmission. APT is also intended to resolve problems associated with the increased flow of patients to services caused by preventable infections and poor PN. It aims to reduce the reinfection and return visits amongst index patients and their sex partners and address problems that relate to barriers to accessing existing sexual health services.

#### Components

APT is multi-agent and multi-level (*see components*) and focuses on three interacting groups of people (i.e., index patients, sex partners and health care professionals) across a range of settings (the sexual health service, the home^2^, telehealth and remote interactions). To deliver APT HCPs are to be trained in how to categorise types of sexual partner, how to facilitate index patients choices of PN approach per sexual partner, how to use a range of behaviour change techniques to educate and persuade index patients to engage with PN and complete their chosen approach. Index patients were offered facilitated discussions of PN approaches for different partner types, they were provided with structured support concerning how to communicate STI diagnoses and encourage PN with their partners and provided with a demonstration of the APT pack and its varied contents. Sex partners were provided with a telephone consultation with an HCP focusing on their choice of testing and treatment and APT delivery (i.e. index patient vs HCP postal). They were talked through the content of the APT pack, offered links to fuller on-line video support for pack use. Then they were provided with a pack compartmentalised to include various self-sampling kits, written instructions on how to use the kits, treatment and a return envelope for completed samples.

#### Mechanisms

We also provide a brief overview of *how* we conceptualise and theorise these intervention elements working (*under mechanisms*). Within the logic model we use the language of the Michie’s COM-B framework (Michie et al., 2011) to provide a high level description of how we assumed the components to work. Within the more detailed initial programme theory (supplementary file 1) we show how we also used normalisation process theory (Murray et al., 2010) and concepts from systems science (e.g. Rutter et al., 2017).

#### Outcomes

In relation to outcomes, Figure 1 shows that APT is intended to lead to increases inpatient choice, sexual health literacy, and the removal of barriers to care. For sexual health services APT is intended to extend service reach by offering new kinds of care, treat sex partners rapidly by removing the need for them to attend overstretched services in remote locations at award times. Together these factors are anticipated to lead to reduced chlamydia reinfection and reduced return visits from patients. Over time, the investment in better PN is expected to lead to fewer patients using the service.

### Research Question 2: What did we learn about the context and contextual dependencies?

The context in which APT and the LUSTRUM trial were implemented are important for interpreting trial results and to considering the potential value of APT as an intervention for the NHS. Process evaluation data did not further illuminate the ‘*amazonisation*’ of culture, or provide any evidence to contest our assumptions about the social patterning of sexual ill health. However it enriched our understanding of the sexual health service system.

Analysis highlighted the sexual health service system was under-resourced and experiencing mounting pressure from various sources (volume of patients, commissioners demands, staff shortages and loss, impoverished physical resources). Services were constantly attempting to adapt to these pressures striving for efficiencies ‘*Routine care has kind of gone off the radar. Because we struggle to cope with the demand for urgent care’* (HCP FG). Adaptions included diverting patients seeking testing away from the services itself (e.g., to on-line self-testing services or high street pharmacies) to enable existing service capacity to focus on treatment. However, problems with meeting patient need for treatment still endured. HCPs stressed the intensity their service provision using terms like ‘*factory*’ or ‘*it can feel like a conveyor belt of people coming in with Chlamydia*’ *(HCP 1:1)*.

Throughout HCP data there was an acute sense of the omnipresent waiting room and the volume of people always waiting to be seen. Thus despite services reorganising to adapt to various pressures there was a constant flow of patients and a clear sense of there being no slack, or even capacity to reflect and plan, within the system. In some places this led to a culture of short-termism, with immediate demands being prioritised through constant ‘firefighting’. Staffing became a limiting factor within the trial ‘*they manage on a day to day basis with how much staff they have’ (HCP 1:1)* and those with expertise in PN had little opportunity to use it because they were needed ‘*to do more nursing bits of their role, like give people injections’ (HCP FG)*.

Capacity to be actively contribute to the trial and deliver the intervention was thus fragile and in some places was negatively affected by any factors adding to the already highly pressurised system. Drawing on concepts from normalisation process theory (NPT) we found that local leadership (‘*contextual integration’*) could affect wider beliefs within services (‘*communal appraisal’* and ‘*relational integration’*) about the value of APT and the wider trial ‘*I think that is a fear amongst staff that if you do this [APT] <u>more</u> it will affect the productivity’ (HCP FG)*. Furthermore across sites HCPs all highlighted the widespread need to duplicate patient records across two, or often three, distinct I.T. systems disincentivising engagement with the trial (in control or intervention phase) and APT.

In summary analysis concerning the context and contextual dependencies highlighted that the sexual health service system was under considerable strain and in a constant state of attempting to adapt to mounting pressures. This led to a highly pressurised environment and a culture of short-termism in some places. There existing resources tended to be prioritised towards treatment rather than prevention and wider care (such as PN). In some sites, the capacity to accommodate the demands of the trial (an upfront investment in resource) were thus limited. In other sites capacity for the trial and the delivery of APT was subject to fluctuation given even minor changes to local circumstances such as staff illness, local leadership, demands of multiple patient records, passwords for IT systems, local approvals of posting medication or shortages in antibiotic packs. APT could be seen as running counter to the prevailing normalised culture of some of the sexual health services it was trialled within which focussed on immediacy, treatment provision and the volume of people using the service.

### Research Question 3: Were the intervention components delivered as intended?

Across the three groups interacting within APT the core components of the intervention were mostly delivered as anticipated. Although there was some evidence of adaption in areas where this was not explicitly encouraged (i.e. support about communication strategies for the index patient).

#### HCPs

As anticipated, all sites did receive the formal training and many also provided local peer-led training. Support from the trial team was delivered consistently. These components were almost universally very well received. Intervention components that failed to be accessible across many sites were the video support to supplement the formal and peer-led training and the intervention manual ‘*[Lead clinician’s] still on chapter two, he’s got it under his bed, you know*’ *(HCP FG)*. In relation to the local implementation plans these were all operationalised as planned and were changed throughout the duration of the trial as anticipated.

#### Index patients

Intervention fidelity for all index patients was generally high. Index patients received facilitated discussions about PN choices per partner and the option of delivering the APT self-sampling and treatment pack to their partner. Postal alternatives were not available in all sites. For index patients choosing APT for their partners, full APT packs were provided with accompanying demonstrations of how to use the pack from HCPs. Support for communication strategies with sex partners were inconsistently delivered in several sites and over the duration of the trial, as in some sites, in order to drive efficiencies and reduce time within the APT interaction itself, index patients were encouraged to tell their sex partner their diagnosis prior to the APT interaction taking place. This also helped to ensure that the sex partner could be available for the telephone consultation with the HCP.

#### Sex partners

Sex partners interviewed all reported having engaged in a telephone based consultation about their choices of PN including APT. Sex partners interviewed reported receiving the APT pack and its return envelope, usually within a day of their consultation. However, no sex partners received links to the instructional videos for remote support with the self-sampling packs. This latter finding was also reported by HCPs in all sites.

### Research Question 4: Is there evidence to support the causal mechanisms by which the intervention was intended to work?

Across the participant groups overall there was a moderate level of support for the putative mechanisms detailed within our initial programme theory and summarised within our logic mode (figure l). The colour coding in Table 2 shows that in general APT worked as anticipated for index patients, sex partners and their partnerships. However, for HCPs and the services they worked in, the anticipated intervention mechanisms sometimes did not gain traction at all, or worked counter-intuitively limiting the reach and functioning of APT.

**Table 1.**
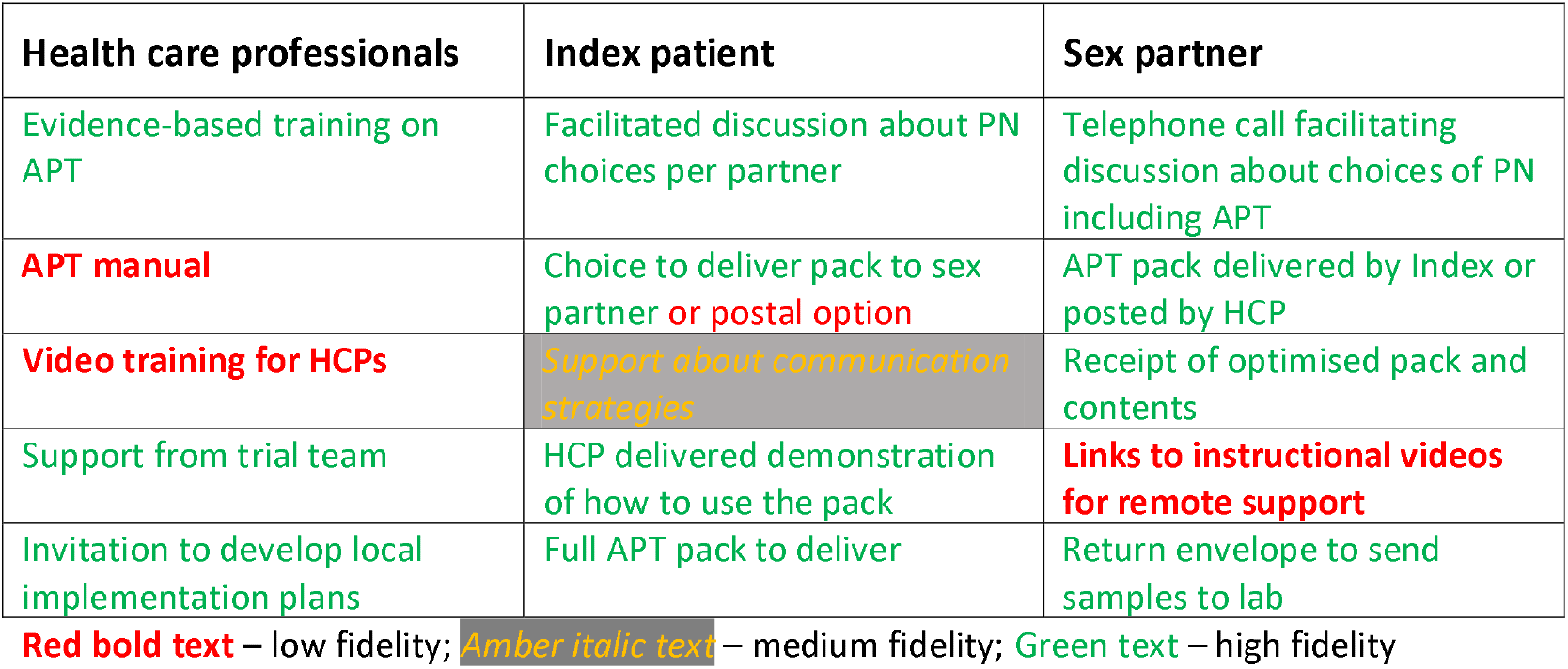
An overview of intervention fidelity and adaptation.

| Health care professionals | Index patient | Sex partner |
| --- | --- | --- |
| Evidence-based training on APT | Facilitated discussion about PN choices per partner | Telephone call facilitating discussion about choices of PN including APT |
| **APT manual** | Choice to deliver pack to sex partner or postal option | APT pack delivered by Index or posted by HCP |
| **Video training for HCPs** | *Support about communication strategies* | Receipt of optimised pack and contents |
| Support from trial team | HCP delivered demonstration of how to use the pack | **Links to instructional videos for remote support** |
| Invitation to develop local implementation plans | Full APT pack to deliver | Return envelope to send samples to lab |

**Red bold text** – low fidelity; *Amber italic text* – medium fidelity; **Green text** – high fidelity

**Table 2.**
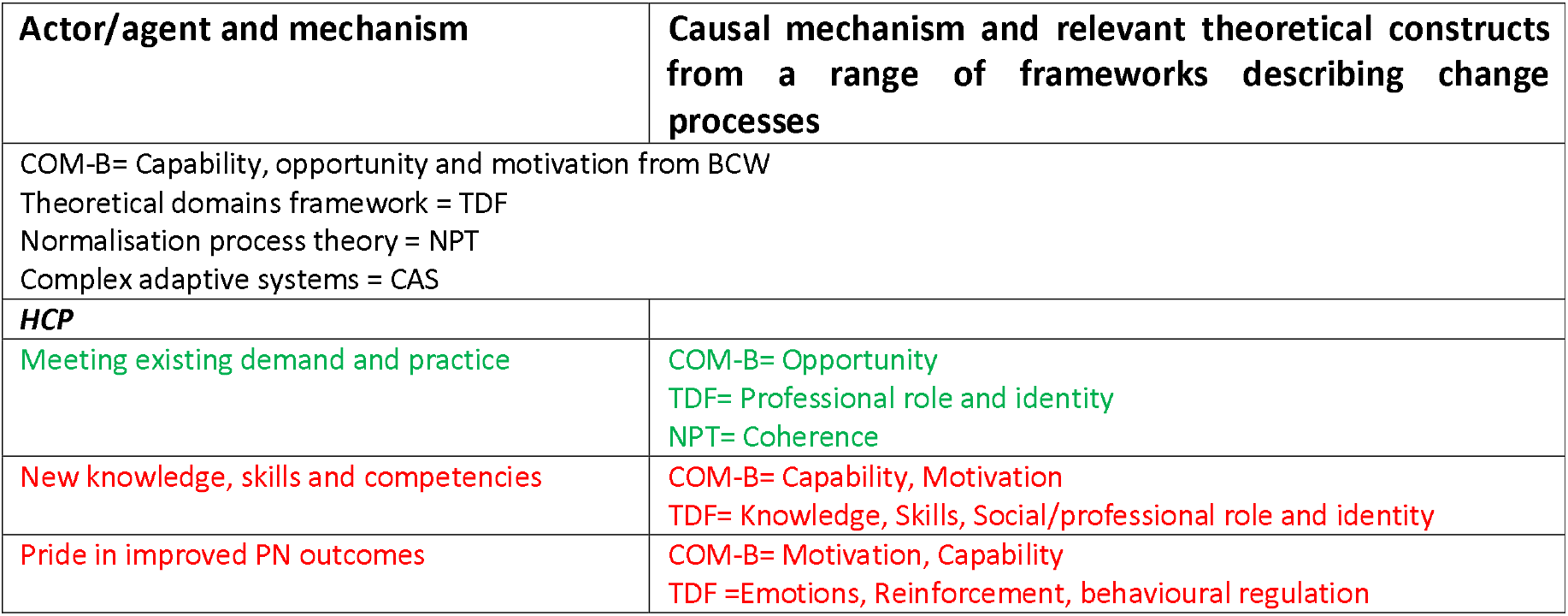
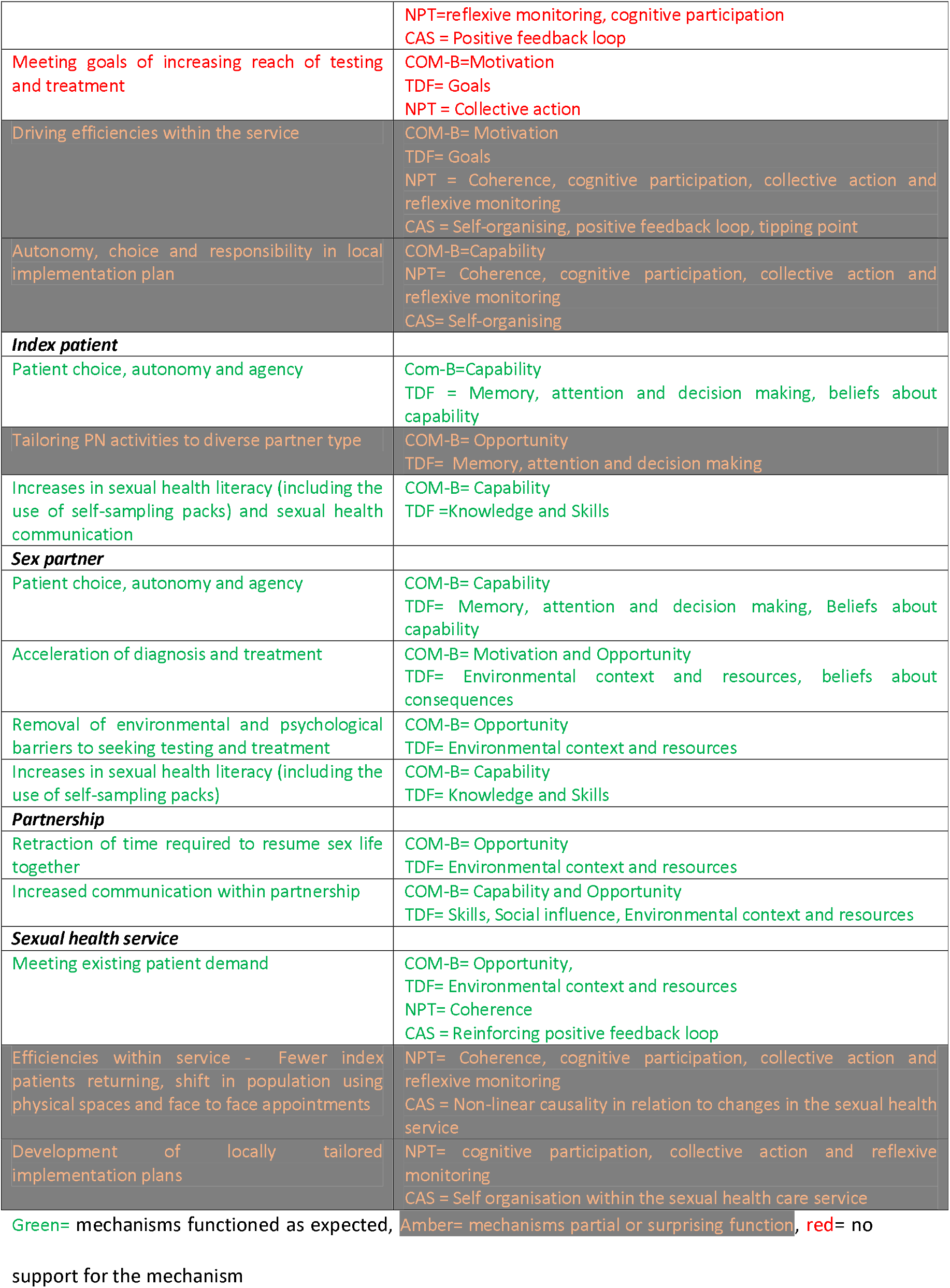
Relative support for the putative casual mechanisms anticipated within APT.

#### HCP

##### Meeting existing demand and practice

There was a clear sense that this mechanism worked as expected. Universally APT was ‘*coherent*’ (May and Finch., 2009) meeting long standing, widespread, patient and provider demand. APT was well liked by those who delivered it.

##### Motivation - for acquisition of new skills, competencies

There was *no* clear support for this mechanism working overall to drive HCPs engagement with and delivery of APT.

##### Motivation - through pride in improved PN outcomes

There was no evidence to support this mechanism working as anticipated. PN outcomes were not mentioned. PN activity did not seem valued within many services and there was no sense of any mechanism by which staff involved with any PN or APT would know of its potential success. This relates to the TDF construct of ‘*behavioural regulation’*, the NPT construct of ‘*reflexive monitoring’* or from systems thinking, the idea of a ‘*positive feedback loop’* enabling the sexual health service system to self-organise in a way that builds on its prior successes.

##### Motivation - through meeting goals to increase reach of testing and treatment

There was no evidence to support this mechanism working as anticipated. It was not a reported goal for individual HCPs or the services in which they worked (NPT’s ‘*collective action’*).

##### Motivation - through driving efficiencies within service

There was no evidence to support this mechanism *enabling* the delivery of APT. Motivation driving efficiencies within the service were central but tended to be to the detriment of APT. Delivering APT was experienced as producing marked inefficiencies within many services. Given its relative novelty it was initially seen as labour intensive and perceived as taking a long time. Particular problems were reported with the apparent invisibility of appointments ‘*the receptionists hated it’* (HCP FG). It appeared as if a single consultation (with the index patient) took an inordinate amount of time and sex partner consultation(s) did not register within service systems at least at the time of the consultation. Moreover given the wider efficiencies APT could deliver were medium and long term, benefits to the HCP and their service were not apparent ‘*I see the principle of it but you don’t see the practice’… ‘the clinic is always going to be full’* (HCP FG). Again there was no way in which a ‘*positive reinforcing feedback*’, or ‘*reflexive monitoring’* could be established and those delivering APT could see its potential effectiveness (i.e. reduction in index patient reinfection and repeat visits):-

> *They were just trying to get through the workload that was in front of them rather than thinking dynamically about it.(p5)’ it’s very easy to feel under pressure if you’ve got a waiting room full of people but sometimes you have to take a step back and say, okay, I have a waiting room full of people but if I take an extra 20 minutes with this person I may save myself 30 minutes tomorrow because their contact won’t be coming into the service’ (HCP 1:1)*

Moreover, short -term efficiencies were understood to benefit sex partners more than anyone else *‘ it’s definitely a benefit for them [sex partner] to just be able to carry on at work and just have a phone call, but obviously, with the clinician, I guess, it’s not saving our time that much’ (HCP 1:1)*. Moreover it could be seen that APT was privileging the sex partners on the phone at home above and beyond people in the waiting room who risked not being seen: ‘*letting those four people down* [in the clinic] *who might have already taken time off work’* (HCP FG). Along these lines APT although rich in ‘*coherence*’ failed to become normalised in some places due to key mechanisms such as ‘*cognitive participation’*, ‘*collective action*’ and ‘*reflexive monitoring*’ being clearly focussed on the wider services rather than APT.

##### Capability - through autonomy, choice and responsibility in developing local implementation plan creating buy-in and delivery

There was strong evidence supporting this mechanism increasing autonomy, choice and responsibility but this did not lead to the effective implementation of APT across most services. The centrality of perceptions that APT exacerbated the existing pressures on services meant that it was sometimes not supported by staff in decision-making roles in some sites ‘*Matron said ‘its just not doable’’* (HCP 1:1). Across the participating sites there was marked heterogeneity in the way APT was offered; delivered by single research nurses or passionate health advisors but typically not becoming normalised across services and offered to all eligible patients within the intervention period. In some sites key mechanisms of normalisation were not attained (e.g. ‘*cognitive participation’*, ‘*collective action’* or ‘reflexive monitoring’).

#### Index patient

##### Capability - through patient choice, autonomy and agency

There was strong evidence to support this mechanism enabling APT as an intervention. Index patients made choices that best suited theirs and their partners circumstances at the time of diagnosis. APT did fail however when sex partners were not available to take part in the telephone consultation when the index patient was with the HCP.

##### Opportunity - through tailored PN to the range of sex partners

This mechanism was partially successful. Tailoring of PN for index patients was often curtailed and APT was not always offered *‘if every single person who was genuinely eligible actually accepted it, for even just one partner, I mean, it would take us forever’* (HCP FG). APT was often only offered for one of the index’s sex partners and this tended to be someone for whom the index had an established relationship with.

##### Capability - through increased sexual health literacy

There is good evidence of index patients increasing their sexual health literacy and facilitating APT processes. Index patients gained sexual health literacy concerning the benefits of effective PN and the treatment of chlamydia. As HCPs taught the index about the contents of the APT pack that they were to take home, index patients also gained knowledge and some practical skills in self-sampling, although not all patients helped their partner complete the self-samples.

#### Sex partner

##### Capability - through patient choice, autonomy and agency

There was strong evidence sex partners benefited from patient choice, autonomy and agency. Treatment rather than testing was a priority for most sex partners. Some wanted to delay their self-sampling until after treatment to make sure they knew the treatment had worked.

##### Motivation and opportunity - through acceleration of diagnosis and treatment

There is strong evidence for APT fast-tracking treatment and diagnosis of sex partners. Sex partners and index patients alike wanted to *‘get rid of it [chlamydia] and move on with my life*.*’* Sex partners commonly received the APT pack within a day of their consultation and most completed the self-samples and took the treatment within a short time frame, giving them quicker access to care than attending a clinic face to face.

##### Opportunity -through the removal of barriers to care

There is very strong evidence that APT removed common barriers to care for sex partners. Many of the sex partners we interviewed described working long, inflexible hours meaning they couldn’t access face-to-face services. The location and opening hours of clinics did not suit everybody and healthcare professionals identified those from rural communities or areas without regular cost-effective public transport as particularly disadvantaged by standard care models.

##### Capability - through increased sexual health literacy

There is good evidence of this mechanism working as anticipated. Sex partners improved their understanding of chlamydia treatment, PN processes and self-sampling procedures. However universally sex partners were not given access to the videos about APT and how to complete the self-samples.

## DISCUSSION

We have used programme theory to structure a process evaluation of APT (a novel approach to contact tracing for chlamydia). Our approach is highly novel using solely qualitative data and combining multi-levelled insights from individual behaviour change, implementation science and complex adaptive systems thinking to give a full and dynamic picture of how APT worked. Using our initial programme theory to make explicit our prior assumptions about what the intervention was, and how we thought it should work through, we then collected in-depth qualitative data and analysed it using its main elements to explore how it actually worked. Using this relatively structured approach, our analysis illustrates the dynamic way that APT appeared to work within the context of the LUSTRUM trial across the diverse trial sites.

Overall it highlights a mixed picture; some components worked well and as anticipated but other components did not work well nor as anticipated. By adopting this structured approach and analysing data through the lens of our prior assumptions we are able to distinguish between the relative contributions of features of the context, content and causal mechanisms that may prove important to interpreting trial outcomes and assessing the practical value of the intervention. This structured approach enables a granular yet multi-levelled approach to considering the dynamic functioning of the intervention and enables us to unpick the respective contribution of the diverse elements of programme theory in ways that are useful for future scale up and/or transferability to other contexts and/or health conditions.

We have used empirical and theoretical lenses to evaluate APT as a complex intervention in the pressured system of sexual health services. We have shown *how* APT was designed to work and detailed the interactions between the context, the problem being addressed, intervention components, key intervention mechanisms and outcomes. Our analysis suggested APT is an acceptable and well liked intervention for some HCPs, IPs and SPs alike. The intervention components were largely delivered as planned to HCPs IPs and SPs. Key mechanisms intended to enhance the offer and receipt of APT worked well and as anticipated. They enabled some HCPs to offer wider choices of PN approaches by partner type to index patients. In turn index patients could choose APT and ensure their sexual partners had further choices of their preferred PN approach. When sex partners chose APT they received an HCP delivered telephone consultation and a self-sampling and treatment pack. Despite previous intervention development work leading to the production and availability of streaming on-line videos providing further detailed support to sex partners to use these packs, universally sex partners were not directed to these intervention resources.

Analysis highlighted problems in some trial sites with delivering both the trial and APT at scale. In order to accommodate the heterogeneity of trial sites and their unique set ups and staff mix and enable local ownership and commitment to the trial and APT, the training and pre-trial work was committed to enable each site to develop local implementation plans. These did not specify exactly how APT was to be offered within given sites, for example, offering dedicated APT clinics within each service, or detailing a minimum number of staff trained and ready to use the trial software and/or to deliver APT. Given the impact of diverse pressures on sexual health services and the perception that APT was inefficient, time consuming and led to no palpable positive outcomes, some sites contracted their offer of APT from across the whole service or APT clinics to a few dedicated staff, or in two sites a dedicated research nurses. Other sites began to offer APT only to those where there was a strong perceived likelihood that it would be accepted (the intervention training and emerging experience of delivering APT showed that it was most likely to be used by those in relationships where sex was likely to occur again or new relationships where STIs could be forgiven and explained). In this way some sexual health services self-organised over time to offer APT to few and to those most likely to choose it. The drive for efficiencies also led to some services to self-organise in ways that minimised inefficiencies within APT. Some services began offering APT within the initial telephone conversations where potential chlamydia diagnosis was discussed in an attempt to increase the likelihood of sex partners being primed and ready for subsequent telephone consultations with HCPs. Whilst these self-organised ‘filters’ emerged to ensure APT could be offered more efficiently they probably also reduced the amount of patients registered for the trial.

In relation to what we have learned about APT within the LUSTRUM trial and how it might shape future service provision, it was well liked by index patients and by sex partners. Furthermore, for the vast majority of health professionals involved in delivering it, it was also found to be rewarding and relatively straightforward. Once people had learned how to deliver it and how to efficiently manage relevant IT systems HCPs enjoyed its immediacy. These findings all suggest strong support for the potential roll out of APT in the future. They stress the acceptability of the intervention itself and its intuitive logic, its coherence and its capacity to be normalised as an additional PN option for sexual health services. Our use of programme theory and its distinctions between context, content and mechanisms has enabled us to see the particular value of the content and many key theorised mechanisms.

However, in contrast to these positive aspects, many of the services in which APT was offered as part of the LUSTRUM trial struggled to integrate it into routine provision within the intervention phase. These issues relate to the context and context-dependencies of APT. Our analysis suggests these difficulties relate primarily to the pressurised context of sexual health services within the UK. Sexual health services were universally described as actively responding to varied mounting pressures and seeking to adapt in order to maximise efficiencies within a dwindling resource envelope (for example offering online self-sampling). Some of these pressures related to the volume of patients seeking sexual health care and some related to the commissioning context and local organisation of sexual health services. In some sites these adaptions (striving for efficiencies) blocked the routinisation and normalisation of both APT and trial procedures. There was a sense that existing resources were all focussed on the immediacy of treatment and very much geared to the palpable volume of people within the waiting room. In these situations APT was seen to be ‘borrowing from Peter to pay Paul’ and the benefits of APT were not immediate and were invisible. These factors do not bode well for the delivery of APT within the future and signal major problems with the delivery of PN and prevention activities overall. They are indicative of a health sector under considerable stress with little capacity for innovation and little tolerance for uncertainty.

Yet in contrast to this somewhat gloomy perspective, our analysis does highlight the centrality of social processes within the sexual health service system and speaks to the potential of harnessing them. Where APT failed to gain traction this was because it was overshadowed by the power of the service system striving to drive efficiencies. It may be possible that evidence of the effectiveness of APT may activate mechanisms such as visible ‘*positive feedback loops’*, NPTs ‘*reflexive monitoring’* or the TDFs ‘*behavioural regulation’*. In this way, evidence of APTs effectiveness may be a ‘*tipping point’* which enables the enormous power of social processes to drive the normalisation of APT within sexual health services if they are seen as contributing to efficiencies. Yet this does not necessarily relate to the results of the LUSTRUM trial but could relate to better systems of monitoring PN outcomes too.

### Strengths and weaknesses of this study

The strengths of the study are its use of programme theory to structure the process evaluation and the triangulation of data sets from different participant groups. In addition, another strength is our use of diverse multi-levelled theoretical lenses to understand the complementary mechanisms at play within the intervention. Other strengths relate to our formulation of initial programme theory.

Weaknesses relate to our reliance on solely qualitative data and analysis and to our recruitment approach which only focused on participants who had delivered or received APT rather than also encompassing those who did not.

## CONCLUSION

We used programme theory to conduct a process evaluation of APT within the context of the LUSTRUM programme. Our analysis showed the different roles of key elements of our programme theory in shaping the implementation of APT within the context of a trial. Our analysis highlighted the centrality of the context and context dependencies and their role in limiting the scalability of APT outwith a trial context. In contrast, our analysis of the content and many key mechanisms of APT supported its acceptability and potential to be normalised should the wider context of sexual health services be changed.

## Supporting information

Supplemental table 1: Initial programme theory

Supplemental file 2: Topic guides

## Data Availability

Data are available upon reasonable request to the study authors

## Footnotes

1 The period of time for which index patients were asked about their sex partner(s) depended on the look-back period (BASHH guidelines) for the diagnosed infection: 3 months for symptomatic chlamydia and 6 months for asymptomatic chlamydia

2 Most participants reported engaging with at least part of the APT process from the place they or their partner lived but this could be any setting away from the physical space of the sexual health clinic

