## Supplemental table 1: Initial programme theory for "Accelerated Partner Therapy contact tracing intervention for people with chlamydia: the LUSTRUM process evaluation using programme theory"

**Supplementary file 1: initial programme theory**

**Initial programme theory of APT**

### The LUSTRUM trial of Accelerated Partner Therapy

The Limiting Undetected Sexually Transmitted infections to RedUce Morbidity (LUSTRUM) research programme aims to improve the sexual health of heterosexual people and men who have sex with men (MSM); the focus is on preventing transmission of sexually transmitted infections (STIs) and reducing undiagnosed HIV through improving partner notification (PN), specifically using Accelerated Partner Therapy (APT). APT links the process of partner notification directly with access to appropriate healthcare by offering sex partners of diagnosed index patients a phone consultation with a healthcare professional who can then arrange delivery of STI testing and treatment via the index patient or by post (see figure xx). Behavioural aspects of self-sampling STI kits were analysed to identify barriers and facilitators to use in order to optimise this the testing component. LUSTRUM builds on previous work which showed APT to be a feasible, acceptable method of partner notification that complied with UK prescribing regulations and resulted in a greater proportion of sex partners receiving treatment (#Estcourt 2012 and 2015). A cluster cross-over randomized controlled trial was conducted in sexual health clinics in England and Scotland to compare the offer of APT to standard PN for people with *C. trachomatis*. The primary outcome was genital C. trachomatis positivity in index patients 12-16 weeks after receiving treatment and initiating partner notification. The full protocol has already been published (#Estcourt 2020).

*Figure 1: APT intervention processes for healthcare professionals, index patients and sex partners*

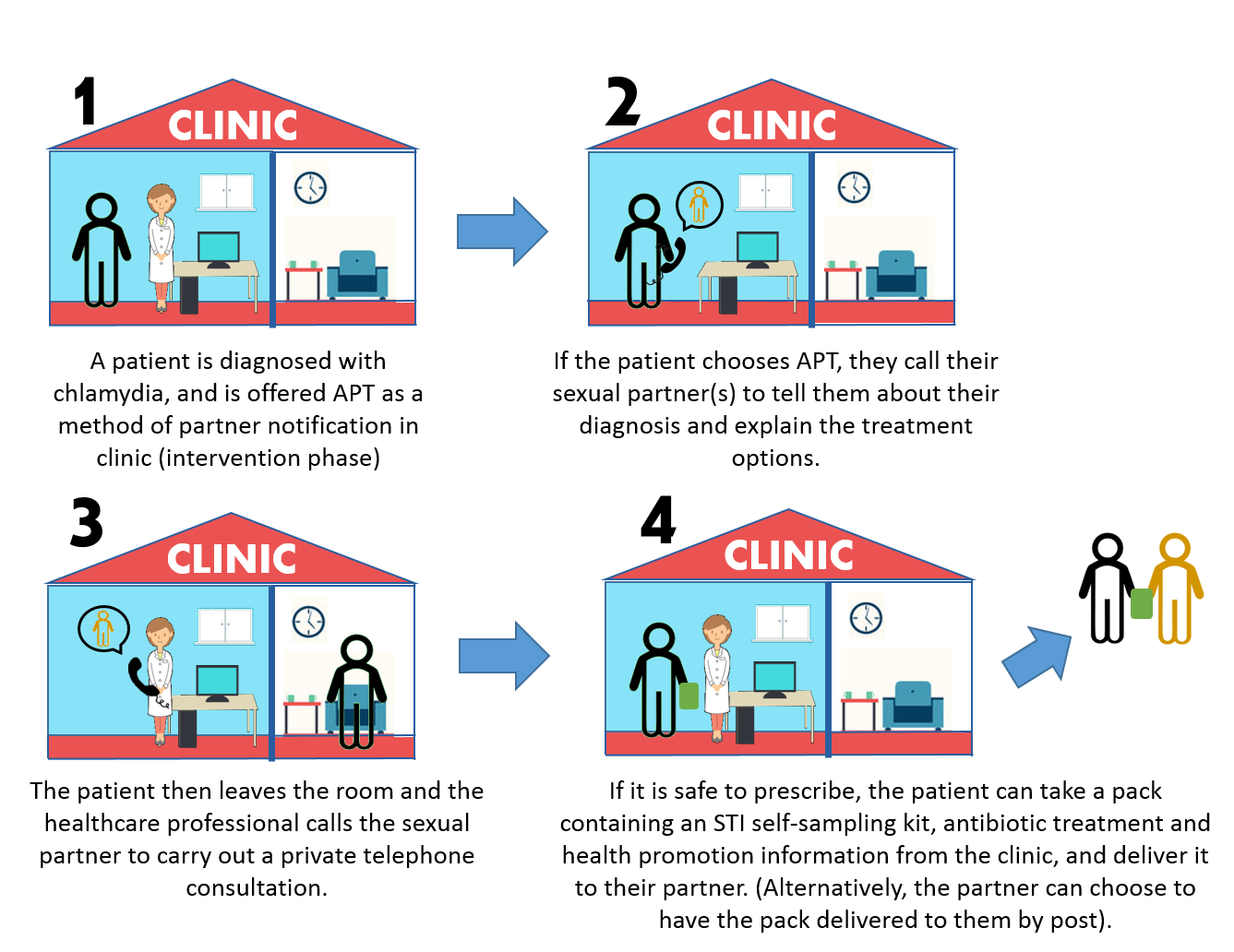

### Partner notification for STI control

Partner notification practices are critical for the management of STIs as they enable sex partners of index patients diagnosed with STIs/HIV to be informed about potential STI risk and to access care (Maclean et al. 2013). PN can help reduce the individuals’ risk of reinfection and STI sequelae and protect wider sexual networks from onward transmission through timely intervention (Bell and Potterat 2011), however it can be challenging. The process relies on the willingness and ability of index patients to identify partners (SHAA PN guidance 2015) however, patients may be resistant due to STI stigma (#stigma refs) and fear of their partners’ reaction to often unexpected information. Partner type is important when considering PN strategies (#Brook 2013 BASHH SH guidelines) and there is some evidence that PN is more common within stable sexual relationships (Braun 2017). APT was developed in response to patient demand for additional options to enable their sex partners to be tested and treated for STIs. It leverages the relationship between index patients and sex partners to provide access to healthcare and exploits advancements in communication technologies and postal self-sampling.

### Theorised change leads to focussed process evaluation

Programme theory is intended to enhance the development and evaluation of complex interventions (MRC, forthcoming). Process evaluations are typically used to assess issues such as the acceptability, fidelity/adaption, to explore the actual functioning of putative causal mechanisms, and to identify contextual factors associated with variation in outcomes (Craig et al., 2008). The key elements of programme theory are variable but Table 1 shows typical content. Critically, programme theory attempts to describe how these elements combine to deliver an effective intervention that addresses the key problem with key intervention solutions within a particular context. By making these clear it is possible to focus process evaluations on collecting information which may be particularly useful for explaining trial results, yet also enabling thorough considerations of issues of future sustainability and transferability at an early point in time.

*Table 1. Typical elements of initial programme theory and how they can structure process evaluation*

| **Key component of programme theory** | **Why include?** | **Key questions to be addressed in developing programme theory** | **Key questions to be addressed within a process evaluation** |
| --- | --- | --- | --- |
| **Context** | Growing recognition of the centrality of *context dependency* in shaping intervention implementation and effectiveness | What assumptions of the context are made to enable the intervention to work?  How might the specificity of the context shape the intervention?  Are concepts from systems science useful for understanding the context in which interventions are embedded? | How did the context of the intervention and the wider trial shape what occurred?  What key elements of the context are central to understanding trial results?  What does an understanding of the context tell us about future sustainability?  What does an understanding of the context tell us about the transferability of the intervention to other contexts? |
| **Problem** | Important to clarify *the nature of the problem* being addressed in order to consider how the intervention components address it and lead to key outcomes | What exactly is the problem being addressed by the intervention? | Did the intervention actually address the problem that was identified?  Was there mission drift or adaption to other emerging problems? |
| **Intervention components** | Important to understand *what* an intervention is made of and how its component parts relate to each other; an understanding of these are central to questions of adaption or fidelity | What is the intervention made of? Who needs to do what to whom, when and in which order?  How do intervention components relate to each other  Are there additive effects?  What are meaningful intervention boundaries? | Were the intervention components delivered as intended (fidelity)?  Were new intervention elements?  Was there local adaption of intervention materials that may have changed the way the intervention worked? |
| **Causal mechanisms** | Important to theorise *how* intervention components actually work separately and in relation to each other. | What are the putative mechanisms by which the intervention components work?  At which levels should we theorise causal mechanisms? Which combination of individual, setting and or system?  How do the putative causal mechanisms interact with each other?  What are the unanticipated mechanisms that shape outcomes? | Is there evidence to support the causal mechanisms by which the intervention was intended to work?  Which mechanisms worked and which did not?  Was there evidence of the intervention mechanisms working together in ways which were not anticipated? |
| **Key outcomes** | What are the key outcomes the intervention is aimed at changing? How are these inter-related | What are the anticipated consequences of the intervention?  What are the unanticipated consequences of the intervention? | Can the process evaluation add value to trial outcomes?  How does a detailed understanding of the intervention enhance the interpretation of trial findings? |
|  |  |  | How acceptable was the intervention? |

**Anticipating the key elements of context and associated context-dependencies**

In relation to understanding the wider context in which the APT intervention was created and delivered, here we address both salient issues of the wider social context and then the contextual dependencies of the trial and the intervention.

***Contexts***

dependent areas; i) The social pattering of sexual ill-health, ii) the ‘amazonisation’ of contemporary culture, and iii) ‘current sexual health commissioning’.

The *social patterning of sexual ill health* is driven largely by the social determinants of health and the cumulative effects of the unequal distribution of power, money and resources (Marmot et al.). Most social determinants of health lie outside of the healthcare system (such as education, working conditions and housing) and are long-standing, intractable issues that, to date, have not been adequately addressed through previous public health, or wider social interventions. These distal factors result in marked social stratification and inequalities in the UK and continue to work against all sexual health improvement efforts. The social patterning of sexual ill-health is therefore embedded in long-standing, complex inequalities, such as the effects of class, racism, homophobia/biphobia/transphobia and a lack of gender equality shaping agency and determining health. Together these distal factors result in a disproportionate sexual health burden for women and several minority groups with the worst effects being realised amongst those with intersecting identities. In turn these determinants of sexual ill-health are also compounded by the social patterning of health and digital literacy and related issues such as sexual health care seeking. Together these social disadvantages intersect with wider social and inter cultural stigmas relating to the transmission and acquisition of STIs, the number of sexual partners people may have or be perceived as having, sex outside of relationship boundaries, and engagement in specific sexual practices. Together this maelstrom of inter-related factors shape observable and long standing epidemiological trends such as the increases in STI incidence and prevalence. Against this backdrop, largely concerned with the distal determinants of sexual ill-health, it is not surprising that many public health interventions have failed to gain traction and that historically, the UK has struggled with effective sexual health promotion. Equally, and particularly pertinent to APT, sexual health services have often struggled to achieve improvements in partner notification outcomes and have been unable or unwilling to prioritise innovation in supporting patients to tell their sex partners about STI diagnoses.

A second key feature of the context is what could be termed the ‘amazonisation’ of lived experiences. This reflects the ubiquity of mobile phones and other digital devices, a growing expectation for real time, immediate, and synchronous communication within many interactions ranging from consumer to health behaviour. It also relates to wider cultural aspects of commodification and a trend towards home-delivery of a range of products. Together these aspects of culture reify and amplify ideas of agency, autonomy and choice within society. Within the health field they have been associated with an increased focus on self-management and self-care. These factors have also shaped, and been shaped, by the ways that sexual cultures are changing and are realised through technology such as mobile phone dating apps. Portable mobile devices can enable the immediacy of sexual interactions, they foster the dehumanisation and reframing of people as products and they disrupt traditional patterns of sexual mixing and add to rising incidence of STIs. Unsurprisingly these aspects of the wider cultural context also shape patient expectations of healthcare. There is a clearer sense of the patient as consumer and the reification of ideas of patient choice and involvement.

The third key feature of the context that is important to consider is the cumulative effect year-on-year of changes in sexual health commissioning and service provision that were implemented as part of the Health and Social Care reform in 2013. In some UK countries, budgets for sexual health services have been transferred away from NHS organisations to local authorities and prevention activities have been split across local and national teams. In this environment private companies can bid against each other and the third sector to provide sexual health services. This has introduced a drive for cost savings and efficiencies and enshrined an outcome culture where the measurement of activities through patient records and other monitoring systems is driven as much by the need to satisfy service level agreements as it is for patient benefit. In turn, commissioning priorities have also shaped wider trends for remote care and self-management. Furthermore, as sexual health service providers are being asked to do more despite significant funding cuts, the drive for efficiencies has been associated with wider trends to change staff skill mix and profession, to reduce staff numbers and to de-prioritise particular areas (such as prevention and partner notification). In light of the epidemiological trends of increasing STI incidence and prevalence it is also noteworthy that more and more patients are trying to access sexual health services and there is growing unmet sexual health need exacerbating existing upward trends in STI prevalence. As a result, healthcare providers report difficulties in conducting effective partner notification and reaching all sex partners of patients who have been diagnosed with an STI. Given the central role of partner notification in reaching people with the highest likelihood of acquiring and transmitting STIs it is clear how this driver also contributes to the increases in STI transmission widely reported across the UK (PHE/HPS/Wales + NI).

***Context dependencies***

There are a number of context dependencies related to the context of STIs and wider sexual health in the UK as described above that are highly salient to APT and the LUSTRUM trial. For the APT intervention, and the wider LUSTRUM trial to work, it is essential that there are sufficient numbers of people with chlamydia who could, and are able to, benefit from a partner notification intervention. It is also essential that sufficient numbers of staff are able to offer APT. The trial and intervention also assumes that people have sufficient health literacy and self-management skills to engage with APT and that they are able to participate in synchronous communication (with their partner and healthcare professionals). Critically, this latter point may intersect with the social patterning of sexual ill-health as those in skilled manual (e.g. within a factory) or highly controlled jobs (e.g. call centre) may have less flexibility to take part in a telephone consultation with an HCP than those with more work autonomy.

Finally, in relation to the provision of sexual health services it is essential that providers have the will and resources to support the trial and delivery of APT as an innovative potential solution to enabling improved partner notification. A core element of provider will and capability is the short term tolerance for trial procedures that may pay off in the long term in relation to a potentially cost-effective, time-saving, pragmatic intervention such as APT that might reduce STI transmission. On a more practical level it is essential that the sexual health services offering the APT and the trial have a series of resources to enable the intervention to be offered and have fidelity (e.g., space, human resource, medication, testing kits, antibiotics, IT systems that enable the sharing of links to videos for staff support for APT and sex partner support for using self-managed testing and treatment packs).

***The specific problems to be addressed by APT***

APT does not address many of the more distal determinants of sexual ill health outlined in the context section above. Its primary focus is on the more proximal determinants of sexual ill-health as they relate to PN.

The primary problem APT addresses is the index patient becoming re-infected with chlamydia because effective partner notification processes including treatment to them and their on-going sexual partners have not taken place. The secondary problem APT aims to address is the delay between an index patient and their sex partner being treated; if their treatment periods do not align (i.e. only one partner is treated) there is an increased likelihood of transmission of untreated chlamydia within the sexual partnership. This is all dependent on the nature of the relationship between the index patient and the sex partner and if they are going to have sex again with each other in the future (#typology paper). To date, partner notification efforts have been poorly targeted according to partner type. These problems relate strongly to some of the contextual issues previously outlined in relation to timely access to testing and treatment and appropriate partner notification approaches being tailored to specific kinds of relationships. Relatedly, and at a much wider scale, APT also makes a modest contribution to addressing the mutually reinforcing problems of high levels of STI transmission, high levels of undiagnosed infections and unmet need in the population discussed above. The way APT is designed and delivered also directly addresses the problem of STI related-stigma; for the sex partner (SP) at least, its remote/telehealth aspects remove many psychosocial and sociocultural barriers concerning STI-related stigma and the related perceived stigma of using sexual health services. In relation to the social patterning of sexual ill-health, APT partially address problems of reach. Because the intervention involves a chain of people connected through mitigating STI risk, the work of the index patient (IP) and healthcare professional (HCP) has the potential to reach those with otherwise low health literacy or the ability to use existing sexual health services. In this way APT has the potential to reach those who may not be reached by other sexual health promotion interventions and resolve the growing problem of sexual health service access and engagement. APT therefore provides a potential means of making a minor contribution to address the social pattering of sexual ill-health. Because APT is intended to engage people with low levels of existing service access and it provides knowledge and some skills APT can also be thought of as having an educational components and that it could enhance levels of sexual health literacy amongst some service users.

At the level of the service, APT is also intended to address many problems associated with the current context of sexual health service provision. In the long term, due to reducing reinfection and onwards transmission, APT is intended to have an impact on reducing the flow of patients in to services. For example, in relation to reducing index patient reinfection, index patient usage of services as a result of reinfection should reduce. It is also possible that higher levels of sexual health literacy in index patients and their sexual partners will also resolve problems of low sexual health awareness. Equally by offering remote care to a range of SPs there will be less use of the physical service. Together these factors may make some contribution to the ongoing problem of rising STI incidence.

***The key intervention elements to be delivered as APT***

The APT intervention itself has three interconnected key actors – the index patient (IP), their sex partner (SP) and the healthcare professional (HCP) bound together in a series of interactions. The intervention takes place within two main settings: the clinic, and the home (or other familiar setting away from the clinic) wherein all actors must work together to enable successful delivery. Each actor receives different intervention components which when used in combination, and through collaboration with the other actors, enable APT to work as a meaningful whole. In this way APT is a complex intervention with multiple inter-dependent components. There is a sense that the intervention must work as a ‘relay’; each person must play their part and keep the baton moving.

*The index patient* receives a facilitated discussion about their sexual history and *the choice* of using APT as one PN option to notify particular partners, specified by partner type. This usually takes place at the time when the index patient has returned in person to a sexual health service to receive treatment. They receive emotional and practical support when contacting their partner from the HCP and can choose for the healthcare professional to be present when initially calling their partners. They may also use suggested communication strategies with their chosen sexual partners (in the form of a checklist provided by the HCP) based on specific behaviour change techniques. If it is safe for them to do so, the index patient may choose to take an APT pack to their sexual partner and help them use it appropriately after tailored explanation of the testing and treatment processes by the HCP; alternatively, the APT pack can be posted by the sexual health service directly to the sex partner.

After *sex partners* have been contacted by the index patient and choose to take part in APT, sex partners receive a phone consultation with a healthcare professional (usually a shortened version of the consultation they would receive if they had attended a clinic) to ensure it is safe for them to receive the APT pack. The pack contains self-sampling STI kits, written instructions, health promotion information and treatment for chlamydia (to be taken after completing the self-samples). They also have access (via a link sent by the HCP) to instructional videos on how to complete the self-samples and a return envelope to send their samples for further diagnostic investigation. The index patient provides emotional and practical support to the sex partner in using the pack.

Relevant *healthcare professionals* receive evidence-based training on how to deliver APT, how to categorise different partner types according to a typology and an evidence-based intervention manual and access to an instructional video about delivering APT. Training and resources are to be provided in advance of patient recruitment starting and training top-ups were offered to some clinics as necessary. Relevant healthcare workers also had use of the RELAY webtool to support partner notification and facilitate data collection for the trial. In addition extensive support from the trial team will be available throughout the implementation of APT which will include a centralised partner notification bureau tasked with arranging follow-up with index patients at two weeks and coordination of the index patient re-test at three months.

For the *sexual health service* sites involved in the trial, given the deep heterogeneity of service configuration, commissioning cycles, relative differences in staff mix, diverse staff competencies and patient-populations, individual sites delivering APT were invited to devise their own implementation plans concerning how to operationalise the trial requirements and the delivery of the APT intervention within the parameters of their training and the delivery of key intervention components. However beyond this it was up to each trial site to devise a locally tailored way of delivering APT. Early work in relation to the selection of sites for the trial had established the basic essentials of space and time requirements, sufficient numbers of eligible patients over a set time period and the identification of local APT champions. Their role was to support the roll out and the sustainability of APT, manage relationships with the trial team, enabling an iterative dialogue concerning questions, concerns, problems and solutions arising during APT implementation.

***The key causal mechanisms by which APT is thought to work***

Table 1 (below) summarises the key causal mechanisms associated with each actor or agent within the intervention. It is important to note that one central mechanism is the connectivity between these actors and agents and their diverse activities. Many mechanisms are interdependent upon others, for example, ‘patient choice’ depends upon HCP ‘skills’ and service configuration ‘efficiencies within service’. Equally, HCP’s ‘pride in PN outcomes’ depends upon the actions of both IP and SPs.

***Index patient***

For the index patient particularly, the intervention is intended to work through increasing patient capability, through enabling patient choice concerning the type of PN and patient autonomy and agency. The IP is encouraged to think about the various partners and make decisions about the type of partner notification most appropriate for each partner type and whether APT would work for them. Through discussion with the HCP, about their diagnosis, partner notification, the option of APT and role of delivering the self-sampling pack the IP increases knowledge concerning sexual health literacy. Through discussions with the HCP and intervention materials (checklist) supporting discussions with SP, some IPs will gain sexual health communication skills. These latter mechanisms also increase patient capability, and motivation, enabling them to engage fully with APT.

***Sex partner***

The intervention works through patient choice concerning the SPs decisions concerning the acceptability of APT for them and in this way draws upon patient autonomy and agency. For the sex partner in particular the intervention works through the acceleration of both diagnosis and treatment. These are facilitated by the IP bringing them, or the HCP sending them, the self-sampling and treatment pack. This central mechanism will remove environmental and psychological barriers to the SP seeking testing and treatment. They can avoid the time it would take to get an appointment with an HCP and they can avoid the felt stigma of using sexual health services. Moreover, if the IP chooses to bring the SP the self-sampling and treatment pack, then they also receive tailored proxy sexual health care and may receive practical and emotional support in using the pack. In this way, this part of APT is intended to work by increasing capability (increasing sexual health literacy and building new skills in relation to the use of self-sampling packs), increasing motivation (quicker return to sex life) and increasing opportunities (removing barriers to treatment).

***Partnership (both IP and SP)***

The intervention is also intended to work for the partnership in terms of the retraction of time required to resume sex together. It also works in terms of increasing communication within the partnership concerning sex, sexual health and the relationship itself.

***HCP***

For health care professionals, the intervention is intended to work through increasing HCP capability and motivation through the acquisition of new knowledge, skills and competencies. The intervention is also thought to work by clarifying and valuing the specialist role and skills required to conduct complex partner notification interactions and increasing pride in improved PN outcomes.

***Sexual health service***

For sexual health services, one key mechanism by which the intervention is thought to works is through implicit motivation concerned with meeting existing patient demand. In this way the intervention has coherence for the service and for individual health care professionals involved within delivery. In other words APT is expected to feel like the extension of the usual logic of PN and fit intuitively into patient demand and the practicalities of people’s lives. The intervention also works for the service through extending some of its long-standing and core purposes through increasing service reach in relation to remotely delivered testing and treatment. In this way the intervention is intended to work by reducing re-infections and onwards transmission making positive contributions to public health. In turn this should reinforce the partner notification work taking place within the service. This represents a critical feedback loop in which partner notification success impacts on some aspects of the patient population and the service can actively respond to such changes and offer different or diverse new services. Relatedly, the intervention is intended to work through using local structures and autonomy to encourage services to develop local implementation plans driving the efficient use of resource to the right places (e.g., collective action). The offer of APT drives service reconfiguration to enable those with partner notification expertise and APT training to work with those who may benefit from APT and enables other non-partner notification resources to be targeted elsewhere (i.e., reflexive monitoring) where they are needed most. As APT works in the context of other tailored PN choices for sexual partners there should be reductions in reinfection of index patients and reduced onwards transmission eventually resulting in reduced patient flow to drop in services and reduced return visits.

**Summarising the putative causal mechanisms within APT from a range of frameworks**

Table 2 below draws upon the behaviour change wheel approach (Michie et al., 2011) the theoretical domains framework (Atkins et al., 2017), normalisation process thoery (Murray et al., 2010) and concepts from systems science (e.g. Rutter et al., 2017) to theorise the change mechanism employed.

**Table 2: The putative casual mechanisms used within APT**

| **Actor/agent and mechanism** | **Causal mechanism and relevant theoretical constructs from a range of frameworks** |
| --- | --- |
|  | COM-B= Capability, opportunity and motivation from BCW  Theoretical domains framework = TDF  Normalisation process theory = NPT  Complex adaptive systems = CAS |
| ***HCP*** |  |
| Meeting existing demand and practice | COM-B= Opportunity  TDF= Professional role and identity  NPT= Coherence |
| New knowledge, skills and competencies | COM-B= Capability, Motivation  TDF= Knowledge, Skills, Social/professional role and identity |
| Pride in improved PN outcomes | COM-B= Motivation, Capability  TDF =Emotions, Reinforcement, behavioural regulation  CAS = Positive feedback loop |
| Meeting goals of increasing reach of testing and treatment | COM-B=Motivation  TDF= Goals |
| Driving efficiencies within the service | COM-B= Motivation  TDF= Goals  NPT = Coherence, cognitive participation, collective action and reflexive monitoring  CAS = Self-organising, positive feedback loop, tipping point |
| Autonomy, choice and responsibility in local implementation plan | COM-B=Capability  NPT= Coherence, cognitive participation, collective action and reflexive monitoring  CAS= Self-organising |
| ***Index patient*** |  |
| Patient choice, autonomy and agency | Com-B=Capability  TDF = Memory, attention and decision making, beliefs about capability |
| Tailoring PN activities to diverse partner type | COM-B= Opportunity  TDF= Memory, attention and decision making |
| Increases in sexual health literacy (including the use of self-sampling packs) and sexual health communication | COM-B= Capability  TDF =Knowledge and Skills |
| ***Sex partner*** |  |
| Patient choice, autonomy and agency | COM-B= Capability  TDF= Memory, attention and decision making, Beliefs about capability |
| Acceleration of diagnosis | COM-B= Motivation and Opportunity  TDF= Environmental context and resources, beliefs about consequences |
| Acceleration of treatment | COM-B= Motivation and Opportunity  TDF= Environmental context and resources, beliefs about consequences |
| Removal of environmental and psychological barriers to seeking testing and treatment | COM-B= Opportunity  TDF= Environmental context and resources |
| Increases in sexual health literacy (including the use of self-sampling packs) | COM-B= Capability  TDF= Knowledge and Skills |
| ***Partnership*** |  |
| Retraction of time required to resume sex life together | COM-B= Opportunity  TDF= Environmental context and resources |
| Increased communication within partnership | COM-B= Capability and Opportunity  TDF= Skills, Social influence, Environmental context and resources |
| ***Sexual health service*** |  |
| Meeting existing patient demand | COM-B= Opportunity,  TDF= Environmental context and resources  NPT= Coherence  CAS = Reinforcing positive feedback loop |
| The increased reach of testing and treatment | COM-B= Opportunity,  TDF= Environmental context and resources |
| Efficiencies within service - Fewer index patients returning, shift in population using physical spaces and face to face appointments | NPT= Coherence, cognitive participation, collective action and reflexive monitoring  CAS = Non-linear causality in relation to changes in the sexual health service |
| Contribution to public health | COM-B= Motivation  TDF= beliefs about consequences  CAS = Emergent properties within the wider context in which intervention is embedded |
| Development of locally tailored implementation plans | NPT= cognitive participation, collective action and reflexive monitoring  CAS = Self organisation within the sexual health care service |

***The key anticipated outcomes of APT***

The primary outcome of APT is the reduction of the proportion of index patients who test positive for C. trachomatis 12-16 weeks after they were treated and chose APT as the method of partner notification for their sex partner(s) as a proxy for measuring index patient re-infection. By reducing re-infection rates within sexual partnerships through effective treatment of both the index patient and the sex partner, the incidence of chlamydia infection will decrease leading to reduced chlamydia transmission within sexual networks. Improved access to STI testing via the self-sampling kits in the APT pack for sex partners and the follow-up test after 12 weeks for the index patient, helps diagnose infections more quickly and interrupt potential future transmission of STIs leading to a reduction in STI incidence. Through APT, sex partners gain access to treatment more quickly and a higher proportion of partners are being reached and treated leading to reduced STI sequelae and related health complications, improved sexual health and wellbeing for both index patients and sex partners. Treating partners simultaneously leads to fewer future attendances at sexual health clinics to treat re-infections which maximises the cost effectiveness of this approach and reduced costs of treatment to the sexual health service overall. However treatment of sex partners is heavily patterned by partnership type which in turn is shaped by wider inequalities. This means that undiagnosed STIs are more likely to be concentrated in people with intersecting vulnerabilities and the acquisition of an STI may work to worsen experiences of inequalities particularly in terms of gender and discrimination against non-heterosexual identities. Finally, the APT intervention has established pathways in services for convenient remote access which increases the potential (and actual) reach of that service and opportunities for engagement with sexual healthcare. Remote access removes barriers to accessing services in person and normalises processes related to self-sampling and telehealth which may contribute to the changing expectations of future healthcare beyond sexual health.

#### Outcomes

| **Mechanism** | **Intermediate mechanism** | **Outcome** |
| --- | --- | --- |
| As per diagram | Reduced chlamydia re-infection of index patient | Reduced chlamydia incidence and prevalence (index patient, sex partner)  Reduced chlamydia transmission in sexual network |
|  | Increased access to STI testing (index patient and sex partner) | Reduced STI incidence and prevalence (gonorrhoea, syphilis and HIV) (index patient, sex partner)  Reduced STI transmission in sexual network |
|  | Quicker access to treatment for sex partners  Higher proportion of sex partners treated | Reduced STI sequelae and improved sexual health and wellbeing ( |
|  | Treatment of sex partners patterned by partnership type | STI prevalence patterned by inequalities (?widen)  Undiagnosed infections concentrated in people with intersecting vulnerabilities |
|  | “2 for the price of 1” treating partners simultaneously | Increased cost-effectiveness for testing and treating STIs |
|  | Pathway for convenient remote access;  Increased service reach and engagement  Removal of barriers to accessing sexual healthcare  Familiarisation and normalisation of telehealth and self-sampling | Expectations of future healthcare changed |
