## Supplemental file 2: Topic guides for "Accelerated Partner Therapy contact tracing intervention for people with chlamydia: the LUSTRUM process evaluation using programme theory"

**Supplementary file 2: Topic guides**

**Topic Guide - Focus Groups with Healthcare Professionals**

Introduction

- *Introduction of the facilitator/researcher and participants*
- *Explain information about the study, the purpose of the focus group discussion and the type of questions that will be asked*
- *Explain that the participants are free to leave at any point*
- *Explain the use of the audio-recorder and how data will be handled*
- *Explain what type of information/data will be shared and with whom (anonymity and confidentiality)*
- *Ask participants if they have any questions and take written consent.*

APT experience – Focus Groups with Health Care Professionals

| **Question 1** | **Can you please tell me your professional role in this clinical service?** |
| --- | --- |
| Indicative prompts | What is your professional role?  How long have you been working in this role?  During this time, have you received any training on other PN options/interventions, besides APT? |
| **Question 2** | **What was it like when the clinic was preparing for the trial** |
| Indicative prompts | When the trial started, did you and your colleagues feel prepared? If yes, how?  How was the start of the trial? What were your thoughts at the time?  How did the trial affect your practice? How did the trial affect the running of the clinic in general?  For those of you who received APT training, what was it like?  What did you like most about it? What did you like least about it?  To what extent do you think the training materials helped prepare your for delivering APT? Did you use the Behaviour Change Techniques (BCTs) with your patients? If yes, which ones did you use? Why? |
| **Question 3** | **What has the implementation of APT been like in your service?** |
| Indicative prompts | Can you tell me about all aspects of the APT intervention process that you followed at your clinic?  How do they differ from routine care? How/in what way do these aspects relate to your usual role in the clinic?  What was it like when the clinic entered into LUSTRUM mode/ APT intervention mode? What was it like for you? What was it like for your patients?  What clinic processes and procedures were in place for APT?  For those of you who were actively involved in delivering APT, how did you find delivering APT? What was difficult for you? What was easy for you?  Did the training material relate to how you actually delivered APT? How did you use the intervention manual and things you’ve learned within the training?  How were things ‘translated’ from theory to practice? What were the challenges you faced? How did you overcome these challenges?  Did you experience challenges at a clinic level? What were they? How did the clinic overcome these challenges?  Why do you think some patients chose APT and some did not?  Did some patients chose different PN for different partners? How did you find this?  What other PN options do you offer at your clinic?  Did your clinic provide the postal option for the APT treatment pack?  Overall, what was good about APT while delivering it? What was bad about it? |
| **Question 4** | **Have the wider research procedures for the LUSTRUM trial impacted on the service?** |
| Indicative prompts | How did these procedures interfere with your clinical activities/workload? How did you cope?  Did your clinical service change the way the research procedures/processes were delivered so that they fit in with your clinical setting? How? Can you please indicate what was adjusted and why? |
| **Question 5** | **Has delivering APT made an impact on your practice?** |
| Indicative prompts | Has there been a change in your current practice as a result of the trial/How did you change your practice as a result of the trial? If changes occurred, what were the aspects of your practice that were changed? How? Why? |
| **Question 6** | **Has delivering APT made an impact on the service?** |
| Indicative prompts | How did APT make a difference to the wider functioning of the clinic?  What are the positive effects/outcomes? Who did they affect?  What are the negative effects/outcomes? Who did they affect?  How are they related to the APT intervention?  In which cases have you noticed these effects/outcomes?   - Upon yourself? - With your patients? - With the sex partners of your patients? - With your colleagues? - Across the clinical service/hub? - Across wider health and service issues? |
| **Question 7** | **What do you think of APT as an intervention when used in practice?** |
| Indicative prompts | What do you think works well?  What do you think does not work well?  What did you expect?  How does that compare to your APT training and the manual? |
| **Question 8** | **How is APT compatible with what the clinic does already?** |
| Indicative prompts | What was new/different/challenging for you? |
| **Question 9** | **What – if anything – could be improved in the LUSTRUM/APT intervention?** |
| Indicative prompts | What would you like to see changed? Why do you want to see these changes?  What are the expected benefits? Are the benefits for the patient population? Are the benefits for the clinical service? Are the benefits for you?  Are there any public health benefits by offering APT as a PN option? If yes, what are the benefits? If no, why not? |
| **Question 10** | **Why did your service participate in the trial?** |
| Indicative prompts | What barriers and difficulties were anticipated at a service level? How were these addressed? |
| **Question 11** | **What external/contextual factors might have influenced the trial?** |
| Indicative prompts | What else was happening across the sector that might have influenced patient and clinic *participation* in the trial?  What else was happening across the sector that might have influenced *trial outcomes*?  What else was happening that might have influenced the *implementation*? |
| **Question 12** | **What was it like when your clinic returned to routine care after the delivering APT?** |
| Indicative prompts | How did you find the change?  Did returning to routine care impact on your clinic/workload/activities? How?  Were there any parts of APT that you found useful for routine care/your service? Would you apply them to routine practice/your service? |
| **Question 13** | **Is there anything you feel we may have missed?** |
| Indicative prompts | If yes, can you tell me more about it? |

*Check if participants have any comments regarding the discussion and how they felt about it.*

Thank participants for taking part!

**Topic Guide - Telephone Interview Schedule with Patients/Clinic attenders**

Introduction

- *Introduction of the researcher and participant*
- *Explain information about the study, the purpose of the interview and the type of questions that will be asked*
- *Explain that the participant is free to leave at any point*
- *Explain the use of the audio-recorder and how data will be handled*
- *Explain what type of information/data will be shared and with whom (anonymity and confidentiality)*
- *Ask if participant has any questions and take verbal recorded consent (Audio-recorder on)*

Demographic questions

- *Ask Demographic Questions*

| 1. **Education level** *(please choose the option that best describes your highest level of education)* |
| --- |
| **Primary education** |
| **Secondary education** |
| **College education** |
| **University education** |
| **Other (please specify)** |

| 2. **Relationship status** *(please choose the option that best describes your relationship status, and if applicable, please indicate in your own words how you would describe your relationship status)* |
| --- |
| **Do you currently consider yourself to be in a relationship?** |
| **Yes** |
| **No** |
| **Prefer not to say** |
| **If you are in a relationship, how would you describe your relationship status?**  **(e.g. long-term, dating, cohabiting/living together, short-term/casual)** |

| 3. **Employment status** *(please choose the option that best describes your employment status)* |
| --- |
| **Full-time employed** |
| **Part-time employed** |
| **Self-employed** |
| **Unemployed** |
| **Retired** |
| **Not in employment, education or training** |
| **Other (please specify)** |

APT experience – Index patient

|  | **Index patient** |
| --- | --- |
| **Question 1** | **Tell me about your recent sexual health consultation** |
| Indicative prompts | What led you to visit the sexual health clinic?  What do you remember most about it?  What did you think about the healthcare professional and the way they spoke to you? How did you feel?  What was it like to tell the health care professional about your sexual partners? |
| **Question 2** | **Tell me about the different ways of contacting your sexual partners (partner notification interventions) that were offered to you?** |
| Indicative prompts | Can you tell me about the options the healthcare professional offered you for contacting your partner/partners?  If multiple options for contacting your partners were offered –  How did you make your choice? How did you think they were different? |
| **Question 3** | **What partner notification options/ pathways did you choose? (the way of doing partner notification)** |
| Indicative prompts | Why did you choose Accelerated Partner Therapy (APT) for notifying your partner/partners?  If multiple partner notification ways were chosen, why did you choose these?  Why did you choose different ways for different partners?  What did you think at the time when you made the decisions?  What challenges – if any – did you experience when making these decisions?  Did the healthcare professional help you in making the choice? In what way?  Did the healthcare professional help you in making the phone call to your partner? In what way? |
| **Question 4** | **Tell me about the telephone process** |
| Indicative prompts | Tell me about when you rang your partner. How did it feel? How did it go? What was his/her reaction at the time?  Was the healthcare professional in the room with you when you phoned your partner? How did that make you feel?  What happened afterwards? |
| **Question 5** | **Tell me about when the healthcare professional talked to your partner** |
| Indicative prompts | How did you feel whilst the healthcare professional talked to your partner?  How did it go?  How long did it take? What were your thoughts at the time? |
| **Question 6** | **Tell me about the APT testing and treatment pack** |
|  | Were you given any options for delivering the treatment pack to your partner? (check for face to face OR postal option)  If both options were offered –  Which option did you choose? Why?  What type of information did you receive from the healthcare professional regarding the testing and treatment pack?  Did you get to see what was inside the pack? Did the healthcare professional show you how to the test and treatment are used?  Did you have any questions at that point? How comfortable did you feel to ask questions at that point?  When you left the clinic how did you feel?  Did you feel you had answers to all your questions? If yes, how? If no, what were the questions that you had after you left the clinic?  What were your thoughts? |
| **Question 7 (for face to face delivery option)** | **What was it like to give the APT testing and treatment pack to your partner?** |
| Indicative prompts | When did you give the pack to your partner? (How many days/hours after your visit at the clinic?)  How did you find delivering the APT treatment pack to your partner? How did you feel?  Did you tell your partner how they should use the pack? What did you say?  Did he/she find the treatment pack user friendly? What was most easy about the pack? What was most difficult about the pack? Did he/she follow the instructions in the pack? |
| **Question 8 (for postal option)** | **Tell me about when your partner received the APT testing and treatment pack through post** |
| Indicative prompts | Do you know if your partner received the APT pack? How soon did he/she receive it? How do you think he/she felt?  Did you tell your partner how they should use the pack? What did you say?  Did he/she find the treatment pack user friendly? What was most easy about the pack? What was most difficult about the pack? Did he/she follow the instructions in the pack? |
| **Question 9** | **What has happened since receiving the Accelerated Partner Therapy intervention?** |
| Indicative prompts | In terms of your wider life? (e.g. impact on relationship with partner)  How did you find the instruction of abstaining from sex after treatment? Did you understand the instruction?  Were there any instructions that were most difficult for you to follow? |
| **Question 10** | **How would you describe your experience overall of doing partner notification like this to your friends?** |
| Indicative prompts | What were your expectations (if any) regarding this partner notification option/way of notifying your partner?  Do you think that the APT option is suitable to all people? (e.g. different cultural backgrounds, different relationships).  What are the potential barriers/difficulties (if any) that you think other people may face with Accelerated Partner Therapy?  Would you recommend this way of notifying partners to your friends? If yes, why? What was good about it? If no, why? What was bad about it? What changes (if any) would you suggest?  Would you prefer to have been given different options? If yes, what are they? |
| **Question 11** | **Is there anything you feel we should have talked about but haven’t yet?** |
| Indicative prompts | Any questions? |

*Check if the participant has any comments regarding the interview and how they felt about it.*

Thank participant for taking part!
